## Supplementary information for "Dengue transmission heterogeneity across Indonesia’s archipelago: climate-driven spatiotemporal patterns and policy implications"

2026-02-10

Table 1

### Table S1. Province Reference Index

| Index | Province Name | Admin ID |
|---|---|---|
| 1 | Aceh | 11 |
| 2 | North Sumatra | 12 |
| 3 | West Sumatra | 13 |
| 4 | Riau | 14 |
| 5 | Jambi | 15 |
| 6 | South Sumatra | 16 |
| 7 | Bengkulu | 17 |
| 8 | Lampung | 18 |
| 9 | Bangka-Belitung Islands | 19 |
| 10 | Riau Islands | 21 |
| 11 | Jakarta | 31 |
| 12 | West Java | 32 |
| 13 | Central Java | 33 |
| 14 | Yogyakarta | 34 |
| 15 | East Java | 35 |
| 16 | Banten | 36 |
| 17 | Bali | 51 |
| 18 | West Nusa Tenggara | 52 |
| 19 | East Nusa Tenggara | 53 |
| 20 | West Kalimantan | 61 |
| 21 | Central Kalimantan | 62 |
| 22 | South Kalimantan | 63 |
| 23 | East Kalimantan | 64 |
| 24 | North Kalimantan | 65 |
| 25 | North Sulawesi | 71 |
| 26 | Central Sulawesi | 72 |
| 27 | South Sulawesi | 73 |
| 28 | Southeast Sulawesi | 74 |
| 29 | Gorontalo | 75 |
| 30 | West Sulawesi | 76 |
| 31 | Maluku | 81 |
| 32 | North Maluku | 82 |
| 33 | West Papua | 91 |
| 34 | Papua | 94 |

### 1 Supplementary Tables

#### 1.1 Table S1: Province Reference Index

Province names and their corresponding numerical indices for reference to all maps in the main figures.

Table 2

### Table S2. Annual Dengue Incidence by Epidemic Year

| Epidemic Year[1] | Total Cases | Incidence per 100,000 | Z-score | Year Type |
|---|---|---|---|---|
| 2010-11 | 85668 | 35.68 | -0.53 | Below average |
| 2011-12 | 84716 | 34.83 | -0.58 | Below average |
| 2012-13 | 110821 | 44.97 | 0.01 | Average |
| 2013-14 | 91108 | 36.50 | -0.48 | Average |
| 2014-15 | 138262 | 54.70 | 0.59 | Above average |
| 2015-16 | 185198 | 72.36 | 1.63 | Major outbreak |
| 2016-17 | 101528 | 39.18 | -0.33 | Average |
| 2017-18 | 55794 | 21.27 | -1.38 | Below average |
| 2018-19 | 138751 | 52.27 | 0.44 | Average |
| 2019-20 | 119912 | 44.64 | -0.01 | Average |
| 2020-21 | 54386 | 20.01 | -1.46 | Below average |
| 2021-22 | 118748 | 43.19 | -0.09 | Average |
| 2022-23 | 124250 | 44.67 | 0.00 | Average |
| 2023-24 | 230635 | 81.99 | 2.19 | Major outbreak |

[1]Epidemic year follows July-June pattern to capture continuous outbreak seasons

#### 1.2 Table S2: Annual Dengue Incidence by Epidemic Year

Incidence by July-June annual period and their respective z-scores. Epidemic year “2015-16” represents July 2015 to June 2016. Major outbreak years are defined as z-score > 1 (more than one standard deviation above the mean).

Table 3

### Table S3. Silhouette Statistics (All Provinces)

| Number of Clusters | Silhouette Score |
|---|---|
| 2 | 0.137 |
| 3 | 0.101 |
| 4 | 0.077 |
| 5 | 0.066 |
| 7 | 0.060 |
| 12 | 0.056 |
| 8 | 0.054 |
| 13 | 0.053 |
| 9 | 0.050 |
| 14 | 0.050 |
| 10 | 0.049 |
| 15 | 0.048 |
| 6 | 0.046 |
| 11 | 0.041 |

#### 1.3 Table S3: Silhouette Statistics for Cluster Optimization (All Provinces)

Silhouette statistics for different numbers of clusters (k=2 to k=15) for all 34 provinces. The selected number of clusters (k=12, highlighted) accounts for both silhouette statistics and balance in cluster sizes.

Table 4

### Table S4. Silhouette Statistics (Western Provinces + Kalimantan)

| Number of Clusters | Silhouette Score |
|---|---|
| 2 | 0.125 |
| 3 | 0.098 |
| 10 | 0.081 |
| 9 | 0.081 |
| 11 | 0.078 |
| 8 | 0.074 |
| 13 | 0.072 |
| 12 | 0.071 |
| 14 | 0.067 |
| 6 | 0.066 |
| 4 | 0.066 |
| 5 | 0.062 |
| 7 | 0.060 |
| 15 | 0.060 |

#### 1.4 Table S4: Silhouette Statistics for Cluster Optimization (Western Provinces + Kalimantan)

Silhouette statistics for western provinces including Kalimantan (22 provinces). The selected number of clusters is k=10 (highlighted).

Table 5

### Table S5. Silhouette Statistics (Western Provinces Only)

| Number of Clusters | Silhouette Score |
|---|---|
| 4 | 0.127 |
| 3 | 0.114 |
| 5 | 0.103 |
| 2 | 0.102 |
| 8 | 0.087 |
| 7 | 0.086 |
| 6 | 0.084 |
| 9 | 0.077 |
| 10 | 0.069 |
| 11 | 0.057 |
| 12 | 0.047 |
| 13 | 0.039 |
| 14 | 0.032 |
| 15 | 0.023 |

#### 1.5 Table S5: Silhouette Statistics for Cluster Optimization (Western Provinces Only)

Silhouette statistics for western provinces only (Sumatra and Java-Bali, 17 provinces). The selected number of clusters is k=4 (highlighted), representing the cleanest cluster structure.

Table 6

### Table S6. Province Dissimilarity Within Regions

| Region | Province | Mean DTW | Z-score[1] | Outlier |
|---|---|---|---|---|
| Java & Bali | JAWA TIMUR | 86.77 | 1.76 | Yes |
| Java & Bali | DKI JAKARTA | 85.14 | 0.92 | No |
| Java & Bali | DAERAH ISTIMEWA YOGYAKARTA | 83.29 | -0.03 | No |
| Java & Bali | JAWA TENGAH | 82.70 | -0.32 | No |
| Java & Bali | BANTEN | 82.22 | -0.57 | No |
| Java & Bali | JAWA BARAT | 82.03 | -0.67 | No |
| Java & Bali | BALI | 81.21 | -1.09 | No |
| Kalimantan | KALIMANTAN UTARA | 91.53 | 1.53 | No |
| Kalimantan | KALIMANTAN SELATAN | 85.18 | 0.27 | No |
| Kalimantan | KALIMANTAN TIMUR | 82.59 | -0.25 | No |
| Kalimantan | KALIMANTAN BARAT | 81.99 | -0.37 | No |
| Kalimantan | KALIMANTAN TENGAH | 77.91 | -1.18 | No |
| Sulawesi | SULAWESI BARAT | 114.06 | 1.85 | Yes |
| Sulawesi | GORONTALO | 106.40 | 0.24 | No |
| Sulawesi | SULAWESI TENGAH | 105.08 | -0.03 | No |
| Sulawesi | SULAWESI UTARA | 103.05 | -0.46 | No |
| Sulawesi | SULAWESI TENGGARA | 101.42 | -0.80 | No |
| Sulawesi | SULAWESI SELATAN | 101.37 | -0.81 | No |
| Sumatra | KEPULAUAN RIAU | 106.61 | 1.98 | Yes |
| Sumatra | LAMPUNG | 102.18 | 0.89 | No |
| Sumatra | SUMATRA BARAT | 101.31 | 0.67 | No |
| Sumatra | SUMATRA UTARA | 100.05 | 0.36 | No |
| Sumatra | ACEH | 98.82 | 0.05 | No |
| Sumatra | BENGKULU | 97.15 | -0.36 | No |
| Sumatra | KEPULAUAN BANGKA BELITUNG | 96.70 | -0.47 | No |
| Sumatra | JAMBI | 94.90 | -0.92 | No |
| Sumatra | RIAU | 94.79 | -0.94 | No |
| Sumatra | SUMATRA SELATAN | 93.48 | -1.27 | No |

[1]Z-score calculated within each region. Provinces with $|z| > 2$ flagged as outliers.

#### 1.6 Table S6: Province Dissimilarity Within Regions

Mean DTW distance from each province to other provinces in the same region, with z-scores calculated within each region. Provinces with $|z| > 1.65$ are flagged as temporal outliers whose dynamics deviate substantially from their regional pattern.

Table 7

### Table S7. Regional Climate Index Correlations

| Region | N Years | ONI Correlation | DMI Correlation | Better Index |
|---|---|---|---|---|
| JAVA & BALI | 14 | 0.622 | 0.024 | ONI |
| KALIMANTAN | 14 | 0.903 | 0.508 | ONI |
| MALUKU | 14 | 0.288 | 0.600 | DMI |
| NUSA TENGGARA | 14 | 0.319 | 0.459 | DMI |
| PAPUA | 14 | 0.785 | 0.459 | ONI |
| SULAWESI | 14 | 0.833 | 0.490 | ONI |
| SUMATRA | 14 | 0.675 | 0.512 | ONI |

#### 1.7 Table S7: Regional Correlations Between Climate Indices and Dengue Incidence

Spearman correlations between large-scale climate indices (ONI and DMI) and dengue incidence by geographic region, using epidemic year aggregation (July-June).

### 2 COVID-19 Pandemic Sensitivity Analysis

The COVID-19 pandemic (2020-2024) may disrupt dengue surveillance systems across Indonesia. To assess whether our findings are robust to potential surveillance artifacts during this period, we conducted a sensitivity analysis comparing results from the full study period (2010-2024) with a pre-pandemic subset (2010-2019).

#### 2.1 Methods

We re-ran three key analyses using only pre-pandemic data:

1. **Wavelet phase analysis** to extract province-level dengue timing patterns
2. **Climate-dengue phase coherence** to identify provinces where climate reliably leads dengue
3. **DLNM analysis** for qualifying provinces (coherence  0.85, positive phase lag) to estimate cumulative relative risk

We then compared results across the two periods using three metrics: (1) agreement in phase lag estimates, (2) agreement in phase coherence, and (3) agreement in effect direction (elevated vs. protective) for DLNM-derived cumulative relative risks.

#### 2.2 Results

##### 2.2.1 Table S8: Phase Lag Comparison

Comparison of dengue phase lags (timing relative to other provinces) between full dataset (2010-2024) and pre-pandemic period (2010-2019).

Table 8

### Table S8. Phase Lag Sensitivity to COVID-19 Period[1]

| Prov. | Reg. | Lag (F) | LCI (F) | UCI (F) | Lag (PP) | LCI (PP) | UCI (PP) | Diff | Agree |
|---|---|---|---|---|---|---|---|---|---|
| ACEH | Sumatra | -2.23 | -3.50 | 1.12 | -2.18 | -4.23 | 0.88 | -0.04 | Same direction |
| BENGKULU | Sumatra | 0.56 | -1.17 | 1.79 | 0.08 | -1.23 | 1.74 | 0.47 | Same direction |
| JAMBI | Sumatra | -1.46 | -3.03 | 2.53 | -1.59 | -3.47 | 1.69 | 0.13 | Same direction |
| KEPULAUAN BANGKA BELITUNG | Sumatra | -1.24 | -2.93 | 2.63 | -0.42 | -2.39 | 2.85 | -0.81 | Same direction |
| KEPULAUAN RIAU | Sumatra | -2.41 | -4.40 | -0.41 | -2.78 | -4.40 | 0.11 | 0.37 | Same direction |
| LAMPUNG | Sumatra | -0.25 | -1.67 | 3.12 | 0.07 | -1.90 | 3.27 | -0.32 | Different direction |
| RIAU | Sumatra | -1.93 | -3.82 | 1.67 | -2.18 | -4.22 | 0.89 | 0.25 | Same direction |
| SUMATRA BARAT | Sumatra | -1.03 | -2.26 | 1.62 | -1.26 | -3.17 | 2.02 | 0.22 | Same direction |
| SUMATRA SELATAN | Sumatra | -0.70 | -2.16 | 3.40 | -0.28 | -2.18 | 3.07 | -0.42 | Same direction |
| SUMATRA UTARA | Sumatra | -2.96 | -4.15 | -0.58 | -3.45 | -4.68 | -0.68 | 0.49 | Same direction |
| BALI | Java & Bali | 1.30 | -0.06 | 3.54 | 1.76 | -0.19 | 4.22 | -0.46 | Same direction |
| BANTEN | Java & Bali | 0.79 | -0.62 | 3.72 | 1.47 | -0.43 | 3.96 | -0.68 | Same direction |
| DAERAH ISTIMEWA YOGYAKARTA | Java & Bali | 0.57 | -0.84 | 3.58 | 1.14 | -0.76 | 4.16 | -0.57 | Same direction |
| DKI JAKARTA | Java & Bali | 1.40 | 0.12 | 3.51 | 1.80 | -0.22 | 4.33 | -0.40 | Same direction |
| JAWA BARAT | Java & Bali | 1.33 | -0.13 | 3.56 | 1.74 | -0.28 | 4.23 | -0.41 | Same direction |
| JAWA TENGAH | Java & Bali | 0.53 | -0.88 | 3.70 | 0.96 | -1.00 | 4.04 | -0.43 | Same direction |
| JAWA TIMUR | Java & Bali | 0.29 | -1.28 | 3.62 | 0.54 | -1.43 | 3.74 | -0.25 | Same direction |
| KALIMANTAN BARAT | Kalimantan | -2.09 | -3.45 | 1.31 | -1.85 | -3.68 | 1.43 | -0.24 | Same direction |
| KALIMANTAN SELATAN | Kalimantan | -0.91 | -2.48 | 3.08 | -0.62 | -2.58 | 2.66 | -0.30 | Same direction |
| KALIMANTAN TENGAH | Kalimantan | -0.92 | -2.49 | 3.07 | -0.53 | -2.50 | 2.75 | -0.39 | Same direction |
| KALIMANTAN TIMUR | Kalimantan | 0.26 | -1.30 | 4.17 | 0.80 | -1.17 | 3.92 | -0.53 | Same direction |
| KALIMANTAN UTARA | Kalimantan | 0.86 | -0.53 | 2.12 | 1.69 | -0.19 | 3.97 | -0.82 | Same direction |
| NUSA TENGGARA BARAT | Nusa Tenggara | 0.36 | -1.05 | 4.01 | 0.90 | -1.00 | 4.09 | -0.54 | Same direction |
| NUSA TENGGARA TIMUR | Nusa Tenggara | -0.53 | -1.99 | 3.57 | -0.07 | -1.97 | 3.28 | -0.47 | Same direction |
| GORONTALO | Sulawesi | 0.21 | -1.36 | 3.25 | 0.25 | -1.71 | 3.45 | -0.05 | Same direction |
| SULAWESI BARAT | Sulawesi | -0.37 | -1.25 | 1.75 | -0.64 | -2.52 | 2.64 | 0.26 | Same direction |
| SULAWESI SELATAN | Sulawesi | 0.39 | -0.98 | 3.51 | 0.70 | -1.20 | 3.96 | -0.30 | Same direction |
| SULAWESI TENGAH | Sulawesi | -0.05 | -1.41 | 2.11 | -0.33 | -2.23 | 2.62 | 0.27 | Same direction |
| SULAWESI TENGGARA | Sulawesi | 0.34 | -1.07 | 3.88 | 0.69 | -1.21 | 3.87 | -0.35 | Same direction |
| SULAWESI UTARA | Sulawesi | 0.21 | -1.20 | 3.64 | 0.59 | -1.31 | 3.93 | -0.38 | Same direction |
| MALUKU | Maluku | -0.32 | -1.84 | 2.21 | -0.47 | -2.37 | 2.87 | 0.15 | Same direction |
| MALUKU UTARA | Maluku | 1.03 | -0.25 | 2.37 | 1.43 | 0.20 | 3.35 | -0.40 | Same direction |
| PAPUA | Papua | -0.21 | -1.57 | 2.39 | -0.33 | -2.23 | 2.94 | 0.12 | Same direction |
| PAPUA BARAT | Papua | -0.09 | -1.47 | 1.36 | -0.07 | -1.46 | 1.25 | -0.01 | Same direction |

[1] F = Full period (2010-2024); PP = Pre-pandemic (2010-2019); LCI/UCI = Lower/Upper confidence interval

###### 2.2.2 Table S9: Phase Coherence Comparison

Comparison of climate-dengue phase coherence between full dataset and pre-pandemic period for precipitation and temperature. “Qualified” indicates provinces meeting threshold criteria (coherence  0.85, phase lag  0) for inclusion in DLNM analysis.

Table 9

### Table S9. Phase Coherence Sensitivity to COVID-19 Period[1]

| Prov. | Reg. | Clim. | Lag (F) | Coh (F) | Lag (PP) | Coh (PP) | ΔCoh | ΔLag | Qual (F) | Qual (PP) | Agree |
|---|---|---|---|---|---|---|---|---|---|---|---|
| DKI JAKARTA | Java & Bali | Precipitation | 2.54 | 0.93 | 2.34 | 0.94 | -0.01 | 0.19 | TRUE | TRUE | Same |
| JAWA BARAT | Java & Bali | Precipitation | 2.53 | 0.96 | 2.41 | 0.97 | -0.01 | 0.12 | TRUE | TRUE | Same |
| JAWA TENGAH | Java & Bali | Precipitation | 1.67 | 0.95 | 1.57 | 0.96 | -0.02 | 0.11 | TRUE | TRUE | Same |
| DAERAH ISTIMEWA YOGYAKARTA | Java & Bali | Precipitation | 1.75 | 0.92 | 1.85 | 0.95 | -0.04 | -0.09 | TRUE | TRUE | Same |
| JAWA TIMUR | Java & Bali | Precipitation | 1.18 | 0.95 | 1.06 | 0.98 | -0.04 | 0.13 | TRUE | TRUE | Same |
| BANTEN | Java & Bali | Precipitation | 1.80 | 0.92 | 2.02 | 0.97 | -0.05 | -0.22 | TRUE | TRUE | Same |
| BALI | Java & Bali | Precipitation | 2.52 | 0.96 | 2.47 | 0.95 | 0.00 | 0.05 | TRUE | TRUE | Same |
| KALIMANTAN BARAT | Kalimantan | Precipitation | -0.21 | 0.82 | -0.07 | 0.88 | -0.06 | -0.14 | FALSE | FALSE | Same |
| KALIMANTAN TENGAH | Kalimantan | Precipitation | 0.02 | 0.93 | -0.03 | 0.94 | -0.01 | 0.04 | TRUE | FALSE | Different |
| KALIMANTAN SELATAN | Kalimantan | Precipitation | -0.17 | 0.95 | -0.29 | 0.94 | 0.01 | 0.13 | FALSE | FALSE | Same |
| KALIMANTAN TIMUR | Kalimantan | Precipitation | 0.75 | 0.87 | 0.90 | 0.90 | -0.04 | -0.15 | TRUE | TRUE | Same |
| KALIMANTAN UTARA | Kalimantan | Precipitation | 5.34 | 0.32 | 4.50 | 0.39 | -0.07 | 0.84 | FALSE | FALSE | Same |
| MALUKU | Maluku | Precipitation | -1.11 | 0.73 | -1.64 | 0.72 | 0.01 | 0.53 | FALSE | FALSE | Same |
| MALUKU UTARA | Maluku | Precipitation | -0.52 | 0.66 | 0.10 | 0.69 | -0.03 | -0.62 | FALSE | FALSE | Same |
| NUSA TENGGARA BARAT | Nusa Tenggara | Precipitation | 1.46 | 0.95 | 1.53 | 0.95 | 0.00 | -0.07 | TRUE | TRUE | Same |
| NUSA TENGGARA TIMUR | Nusa Tenggara | Precipitation | 0.41 | 0.95 | 0.56 | 0.96 | -0.02 | -0.15 | TRUE | TRUE | Same |
| PAPUA BARAT | Papua | Precipitation | -0.20 | 0.58 | -0.08 | 0.52 | 0.06 | -0.12 | FALSE | FALSE | Same |
| PAPUA | Papua | Precipitation | 0.15 | 0.64 | -0.72 | 0.65 | 0.00 | 0.87 | FALSE | FALSE | Same |
| SULAWESI UTARA | Sulawesi | Precipitation | -0.84 | 0.83 | -0.70 | 0.85 | -0.02 | -0.14 | FALSE | FALSE | Same |
| SULAWESI TENGAH | Sulawesi | Precipitation | -2.72 | 0.59 | -2.90 | 0.52 | 0.07 | 0.18 | FALSE | FALSE | Same |
| SULAWESI SELATAN | Sulawesi | Precipitation | 0.83 | 0.91 | 0.60 | 0.93 | -0.02 | 0.24 | TRUE | TRUE | Same |
| SULAWESI TENGGARA | Sulawesi | Precipitation | -0.26 | 0.91 | -0.26 | 0.89 | 0.02 | 0.01 | FALSE | FALSE | Same |
| GORONTALO | Sulawesi | Precipitation | -0.67 | 0.76 | -0.31 | 0.83 | -0.08 | -0.37 | FALSE | FALSE | Same |
| SULAWESI BARAT | Sulawesi | Precipitation | -0.56 | 0.59 | -0.35 | 0.74 | -0.15 | -0.22 | FALSE | FALSE | Same |
| ACEH | Sumatra | Precipitation | 0.37 | 0.79 | 0.32 | 0.95 | -0.16 | 0.05 | FALSE | TRUE | Different |
| SUMATRA UTARA | Sumatra | Precipitation | -0.92 | 0.87 | -0.74 | 0.93 | -0.06 | -0.18 | FALSE | FALSE | Same |
| SUMATRA BARAT | Sumatra | Precipitation | 1.76 | 0.73 | 1.42 | 0.88 | -0.15 | 0.34 | FALSE | TRUE | Different |
| RIAU | Sumatra | Precipitation | -0.11 | 0.81 | -0.40 | 0.91 | -0.10 | 0.29 | FALSE | FALSE | Same |
| JAMBI | Sumatra | Precipitation | -0.11 | 0.91 | -0.41 | 0.93 | -0.01 | 0.30 | FALSE | FALSE | Same |
| SUMATRA SELATAN | Sumatra | Precipitation | 0.21 | 0.96 | 0.31 | 0.97 | -0.01 | -0.10 | TRUE | TRUE | Same |
| BENGKULU | Sumatra | Precipitation | 1.67 | 0.59 | 2.14 | 0.47 | 0.11 | -0.46 | FALSE | FALSE | Same |
| LAMPUNG | Sumatra | Precipitation | 0.46 | 0.90 | 0.29 | 0.92 | -0.02 | 0.17 | TRUE | TRUE | Same |
| KEPULAUAN BANGKA BELITUNG | Sumatra | Precipitation | -0.45 | 0.88 | -0.06 | 0.94 | -0.06 | -0.39 | FALSE | FALSE | Same |
| KEPULAUAN RIAU | Sumatra | Precipitation | -0.58 | 0.79 | -0.75 | 0.77 | 0.01 | 0.18 | FALSE | FALSE | Same |
| DKI JAKARTA | Java & Bali | Temperature | -4.41 | 0.94 | -4.49 | 0.92 | 0.01 | 0.08 | FALSE | FALSE | Same |
| JAWA BARAT | Java & Bali | Temperature | -4.72 | 0.88 | -4.71 | 0.86 | 0.02 | -0.01 | FALSE | FALSE | Same |
| JAWA TENGAH | Java & Bali | Temperature | -5.98 | 0.09 | 5.86 | 0.17 | -0.08 | -11.84 | FALSE | FALSE | Same |
| DAERAH ISTIMEWA YOGYAKARTA | Java & Bali | Temperature | 0.84 | 0.82 | 1.05 | 0.90 | -0.08 | -0.20 | FALSE | TRUE | Different |
| JAWA TIMUR | Java & Bali | Temperature | 4.03 | 0.92 | 3.79 | 0.93 | -0.01 | 0.24 | TRUE | TRUE | Same |
| BANTEN | Java & Bali | Temperature | -5.24 | 0.37 | -4.90 | 0.88 | -0.51 | -0.34 | FALSE | FALSE | Same |
| BALI | Java & Bali | Temperature | 2.16 | 0.91 | 2.17 | 0.93 | -0.02 | -0.01 | TRUE | TRUE | Same |
| KALIMANTAN BARAT | Kalimantan | Temperature | 5.57 | 0.14 | 5.87 | 0.11 | 0.03 | -0.30 | FALSE | FALSE | Same |
| KALIMANTAN TENGAH | Kalimantan | Temperature | 5.43 | 0.34 | 5.20 | 0.45 | -0.11 | 0.22 | FALSE | FALSE | Same |
| KALIMANTAN SELATAN | Kalimantan | Temperature | 4.98 | 0.40 | 4.74 | 0.51 | -0.11 | 0.24 | FALSE | FALSE | Same |
| KALIMANTAN TIMUR | Kalimantan | Temperature | 5.97 | 0.25 | -5.32 | 0.21 | 0.04 | 11.29 | FALSE | FALSE | Same |
| KALIMANTAN UTARA | Kalimantan | Temperature | -2.31 | 0.54 | -2.15 | 0.86 | -0.33 | -0.16 | FALSE | FALSE | Same |
| MALUKU | Maluku | Temperature | 1.10 | 0.75 | 0.47 | 0.81 | -0.05 | 0.63 | FALSE | FALSE | Same |
| MALUKU UTARA | Maluku | Temperature | 1.70 | 0.51 | 1.54 | 0.45 | 0.06 | 0.17 | FALSE | FALSE | Same |
| NUSA TENGGARA BARAT | Nusa Tenggara | Temperature | 2.69 | 0.91 | 2.67 | 0.91 | 0.00 | 0.02 | TRUE | TRUE | Same |
| NUSA TENGGARA TIMUR | Nusa Tenggara | Temperature | 1.97 | 0.93 | 1.95 | 0.95 | -0.02 | 0.02 | TRUE | TRUE | Same |
| PAPUA BARAT | Papua | Temperature | 1.76 | 0.56 | 1.28 | 0.53 | 0.04 | 0.48 | FALSE | FALSE | Same |
| PAPUA | Papua | Temperature | 1.58 | 0.67 | 0.80 | 0.65 | 0.02 | 0.78 | FALSE | FALSE | Same |
| SULAWESI UTARA | Sulawesi | Temperature | 1.52 | 0.70 | 0.96 | 0.72 | -0.03 | 0.56 | FALSE | FALSE | Same |
| SULAWESI TENGAH | Sulawesi | Temperature | 1.55 | 0.69 | 0.84 | 0.71 | -0.02 | 0.71 | FALSE | FALSE | Same |
| SULAWESI SELATAN | Sulawesi | Temperature | 3.57 | 0.90 | 3.40 | 0.89 | 0.01 | 0.16 | TRUE | TRUE | Same |
| SULAWESI TENGGARA | Sulawesi | Temperature | 2.45 | 0.85 | 2.46 | 0.82 | 0.04 | -0.01 | TRUE | FALSE | Different |
| GORONTALO | Sulawesi | Temperature | 3.92 | 0.60 | 4.18 | 0.66 | -0.06 | -0.26 | FALSE | FALSE | Same |
| SULAWESI BARAT | Sulawesi | Temperature | 0.25 | 0.54 | 0.58 | 0.65 | -0.10 | -0.32 | FALSE | FALSE | Same |
| ACEH | Sumatra | Temperature | 5.78 | 0.20 | 5.82 | 0.17 | 0.03 | -0.04 | FALSE | FALSE | Same |
| SUMATRA UTARA | Sumatra | Temperature | 4.27 | 0.88 | 4.52 | 0.92 | -0.04 | -0.25 | TRUE | TRUE | Same |
| SUMATRA BARAT | Sumatra | Temperature | -4.01 | 0.50 | -4.27 | 0.87 | -0.38 | 0.27 | FALSE | FALSE | Same |
| RIAU | Sumatra | Temperature | 5.77 | 0.16 | 5.45 | 0.26 | -0.11 | 0.31 | FALSE | FALSE | Same |
| JAMBI | Sumatra | Temperature | 5.87 | 0.09 | 5.51 | 0.23 | -0.14 | 0.36 | FALSE | FALSE | Same |
| SUMATRA SELATAN | Sumatra | Temperature | 5.36 | 0.41 | 5.52 | 0.30 | 0.11 | -0.16 | FALSE | FALSE | Same |
| BENGKULU | Sumatra | Temperature | -1.89 | 0.62 | -2.84 | 0.62 | 0.00 | 0.95 | FALSE | FALSE | Same |
| LAMPUNG | Sumatra | Temperature | 5.35 | 0.49 | 5.39 | 0.52 | -0.03 | -0.04 | FALSE | FALSE | Same |
| KEPULAUAN BANGKA BELITUNG | Sumatra | Temperature | -5.96 | 0.30 | -5.65 | 0.28 | 0.01 | -0.31 | FALSE | FALSE | Same |
| KEPULAUAN RIAU | Sumatra | Temperature | 4.01 | 0.50 | 4.43 | 0.39 | 0.11 | -0.42 | FALSE | FALSE | Same |

[1] F = Full (2010-2024); PP = Pre-pandemic (2010-2019); Coh = Coherence; Qual = Qualified for DLNM

##### 2.2.3 Table S10: DLNM Cumulative Relative Risk Comparison

Comparison of DLNM-derived cumulative relative risks at wavelet-derived lags between full dataset and pre-pandemic period. Only provinces qualifying in the full period are shown.

Table 10

### Table S10. DLNM Relative Risk Sensitivity to COVID-19 Period[1]

| Prov. | Clim. | L(F) | RR(F) | RRL(F) | RRH(F) | Sig(F) | L(PP) | RR(PP) | RRL(PP) | RRH(PP) | Sig(PP) | ΔRR | Both Sig | Dir Agree |
|---|---|---|---|---|---|---|---|---|---|---|---|---|---|---|
| BALI | Precipitation | 3 | 1.18 | 0.72 | 1.94 | No | 2 | 1.26 | 0.71 | 2.23 | No | -0.07 | TRUE | Same direction |
| BANTEN | Precipitation | 2 | 1.13 | 0.77 | 1.66 | No | 2 | 1.92 | 1.40 | 2.64 | Yes | -0.79 | FALSE | Same direction |
| DAERAH ISTIMEWA YOGYAKARTA | Precipitation | 2 | 1.59 | 1.01 | 2.48 | Yes | 2 | 0.67 | 0.45 | 0.99 | Yes | 0.91 | TRUE | Different direction |
| DKI JAKARTA | Precipitation | 3 | 1.59 | 1.04 | 2.43 | Yes | 2 | 2.65 | 1.81 | 3.89 | Yes | -1.07 | TRUE | Same direction |
| JAMBI | Precipitation | 0 | 1.28 | 1.01 | 1.62 | Yes | 0 | 1.12 | 0.86 | 1.46 | No | 0.16 | FALSE | Same direction |
| JAWA BARAT | Precipitation | 3 | 1.15 | 0.78 | 1.70 | No | 2 | 0.84 | 0.61 | 1.14 | No | 0.32 | TRUE | Different direction |
| JAWA TENGAH | Precipitation | 2 | 1.33 | 0.88 | 2.02 | No | 2 | 0.65 | 0.40 | 1.04 | No | 0.69 | TRUE | Different direction |
| JAWA TIMUR | Precipitation | 1 | 0.94 | 0.68 | 1.29 | No | 1 | 0.62 | 0.45 | 0.84 | Yes | 0.32 | FALSE | Same direction |
| KALIMANTAN SELATAN | Precipitation | 0 | 1.48 | 1.14 | 1.92 | Yes | 0 | 1.35 | 1.04 | 1.76 | Yes | 0.13 | TRUE | Same direction |
| KALIMANTAN TENGAH | Precipitation | 0 | 1.65 | 1.37 | 1.99 | Yes | 0 | 1.15 | 0.94 | 1.40 | No | 0.51 | FALSE | Same direction |
| KALIMANTAN TIMUR | Precipitation | 1 | 1.28 | 1.09 | 1.51 | Yes | 1 | 1.22 | 0.96 | 1.55 | No | 0.06 | FALSE | Same direction |
| KEPULAUAN BANGKA BELITUNG | Precipitation | 0 | 1.46 | 1.21 | 1.76 | Yes | 0 | 1.46 | 1.17 | 1.82 | Yes | 0.00 | TRUE | Same direction |
| LAMPUNG | Precipitation | 0 | 1.30 | 1.06 | 1.59 | Yes | 0 | 1.08 | 0.89 | 1.31 | No | 0.22 | FALSE | Same direction |
| NUSA TENGGARA BARAT | Precipitation | 1 | 1.95 | 1.38 | 2.75 | Yes | 2 | 2.30 | 1.09 | 4.84 | Yes | -0.35 | TRUE | Same direction |
| NUSA TENGGARA TIMUR | Precipitation | 0 | 1.60 | 0.95 | 2.69 | No | 1 | 5.58 | 1.93 | 16.12 | Yes | -3.98 | FALSE | Same direction |
| SULAWESI SELATAN | Precipitation | 1 | 1.33 | 1.05 | 1.67 | Yes | 1 | 1.10 | 0.84 | 1.43 | No | 0.23 | FALSE | Same direction |
| SULAWESI TENGGARA | Precipitation | 0 | 1.12 | 0.94 | 1.35 | No | 0 | 1.20 | 0.98 | 1.47 | No | -0.08 | TRUE | Same direction |
| SUMATRA SELATAN | Precipitation | 0 | 1.21 | 1.06 | 1.38 | Yes | 0 | 1.14 | 0.98 | 1.33 | No | 0.06 | FALSE | Same direction |
| BALI | Temperature | 2 | 0.85 | 0.62 | 1.18 | No | 2 | 1.37 | 0.68 | 2.75 | No | -0.51 | TRUE | Different direction |
| JAWA TIMUR | Temperature | 4 | 1.24 | 0.83 | 1.85 | No | 4 | 11.79 | 5.90 | 23.56 | Yes | -10.55 | FALSE | Same direction |
| NUSA TENGGARA BARAT | Temperature | 3 | 0.60 | 0.39 | 0.93 | Yes | 3 | 1.48 | 0.71 | 3.06 | No | -0.88 | FALSE | Different direction |
| NUSA TENGGARA TIMUR | Temperature | 2 | 1.49 | 0.85 | 2.60 | No | 2 | 2.13 | 0.99 | 4.55 | No | -0.64 | TRUE | Same direction |
| SULAWESI SELATAN | Temperature | 4 | 3.36 | 1.71 | 6.61 | Yes | 3 | 0.81 | 0.39 | 1.68 | No | 2.55 | FALSE | Different direction |
| SULAWESI TENGGARA | Temperature | 2 | 1.12 | 0.75 | 1.66 | No | 2 | 0.48 | 0.26 | 0.90 | Yes | 0.63 | FALSE | Different direction |

[1]F = Full (2010-2024); PP = Pre-pandemic (2010-2019); L = Lag; RRL/RRH = RR Lower/Higher CI; Sig = Significant

##### 2.3 Interpretation

The sensitivity analysis shows that our key findings are **robust to the inclusion of pandemic years**. Phase lag directions remain consistent for the majority of provinces, and provinces qualifying for DLNM analysis show substantial overlap between periods. While absolute values of phase coherence and relative risk estimates differ slightly—expected given the shorter time series in the pre-pandemic period—the qualitative conclusions regarding which provinces show reliable climate-dengue timing relationships and elevated dengue risk remain unchanged.

This robustness suggests that the temporal patterns we identify reflect genuine epidemiological dynamics rather than surveillance artifacts, though we acknowledge that case detection likely declined during 2020-2021 and subsequent recovery may have introduced noise.

Table 11

### Table S11. Sensitivity of Results to Phase Coherence Threshold[1]

| threshold | clim_var | n | n_significant | pct_significant |
|---|---|---|---|---|
| 0.70 | Precipitation | 22 | 12 | 54.5 |
| 0.70 | Temperature | 9 | 3 | 33.3 |
| 0.75 | Precipitation | 21 | 11 | 52.4 |
| 0.75 | Temperature | 9 | 3 | 33.3 |
| 0.80 | Precipitation | 20 | 11 | 55.0 |
| 0.80 | Temperature | 8 | 3 | 37.5 |
| 0.85 | Precipitation | 18 | 11 | 61.1 |
| 0.85 | Temperature | 7 | 3 | 42.9 |
| 0.90 | Precipitation | 16 | 9 | 56.2 |
| 0.90 | Temperature | 5 | 2 | 40.0 |

[1]Bold row indicates threshold used in main analysis (0.85)

#### 3 Phase Coherence Threshold Sensitivity Analysis

Our main analysis uses a phase coherence threshold of 0.85 to identify provinces where climate variables show reliable timing relationships with dengue. This threshold represents a high degree of consistency in the climate-dengue phase relationship. To assess whether our conclusions are sensitive to this threshold choice, we tested alternative thresholds ranging from 0.70 to 0.90.

##### 3.1 Methods

For each threshold value (0.70, 0.75, 0.80, 0.85, 0.90), we:

1. Identified provinces meeting the threshold criteria (coherence  threshold AND phase lag  0)
2. Counted qualifying provinces for precipitation and temperature
3. Determined how many qualifying provinces showed statistically significant DLNM effects (cumulative RR confidence interval excluding 1.0)

Higher thresholds are more stringent, selecting only provinces with very consistent climate-dengue timing. Lower thresholds are more permissive but may include provinces where the relationship is less reliable.

#### 3.2 Results

##### 3.2.1 Table S11: Threshold Sensitivity Analysis

#### 3.3 Interpretation

The number of qualifying provinces decreases predictably as the threshold becomes more stringent. The table shows that stricter coherence requirements substantially reduce the number of qualifying provinces, but the number showing statistically significant DLNM effects remains relatively stable, particularly for the 0.75-0.85 range.

Importantly, **the number of provinces showing statistically significant DLNM effects remains relatively stable** across thresholds. This indicates that our choice of 0.85 is conservative but does not substantially affect the core finding: provinces with reliable climate-dengue timing also tend to show significant dose-response relationships in DLNM models.

The 0.85 threshold balances two considerations: (1) not excluding too many provinces for meaningful interpretation, and (2) maintaining high confidence in the reliability of the climate-dengue timing relationship.

### 4 Supplementary Figures

#### 4.1 Figure S1: Geographical Regions of Indonesia

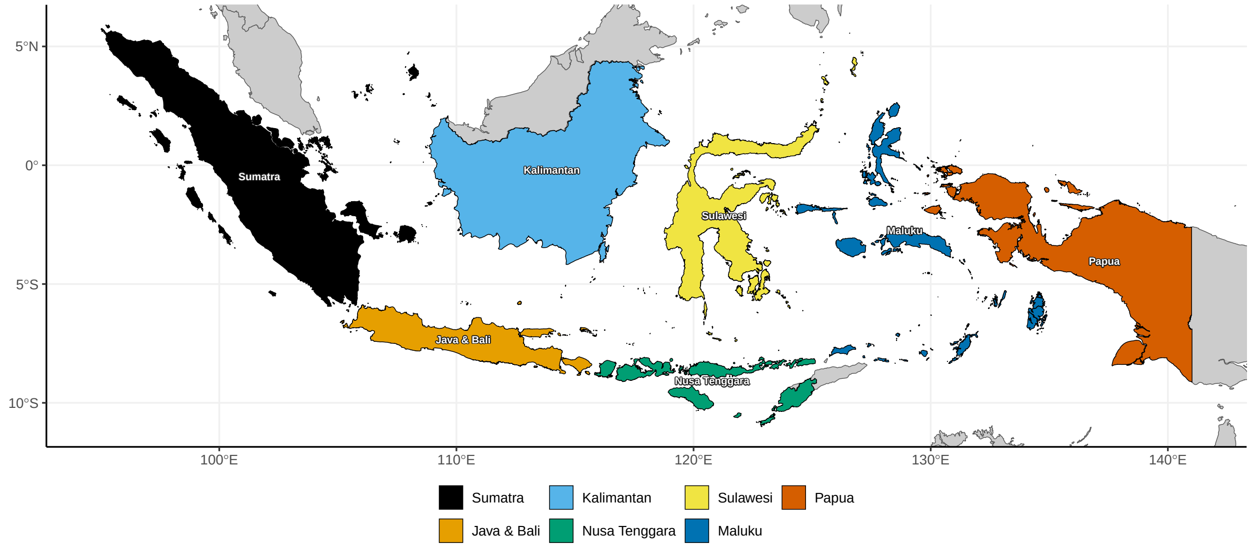

Figure 1: Geographical regions of Indonesia used for regional stratification in this study. Regions follow standard Indonesian geographic divisions.

#### 4.2 Figure S2: Province Phase Lags with Confidence Intervals

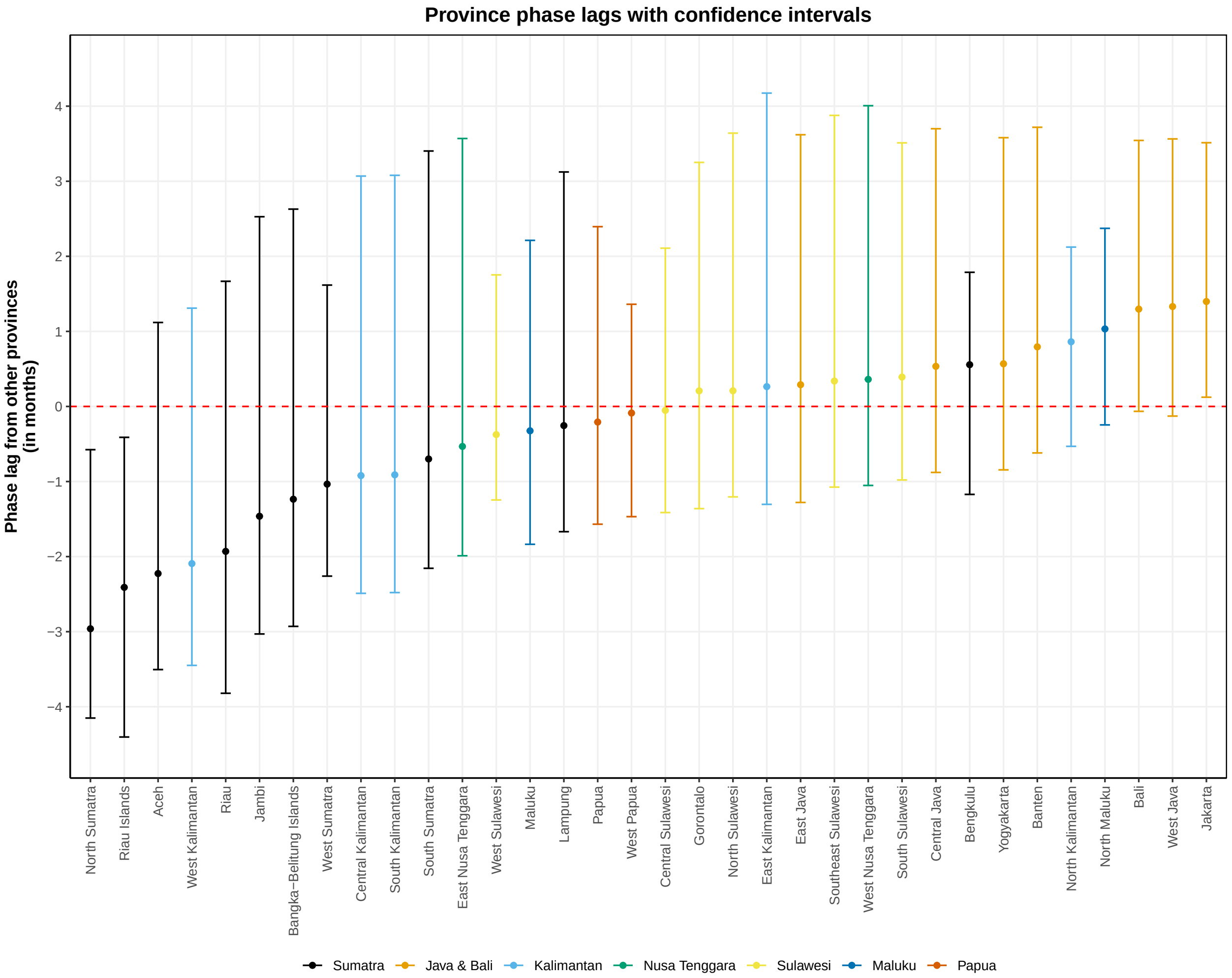

Figure 2: Between-province phase lags with 2.5th-97.5th percentile ranges of pairwise differences. Positive values indicate provinces with later peaks relative to other provinces; negative values indicate earlier peaks. Error bars represent the range of phase differences with all other provinces, not statistical uncertainty.

#### 4.3 Figure S3: Comparison of Wavelet-Based and DLNM-Based Lag Estimates

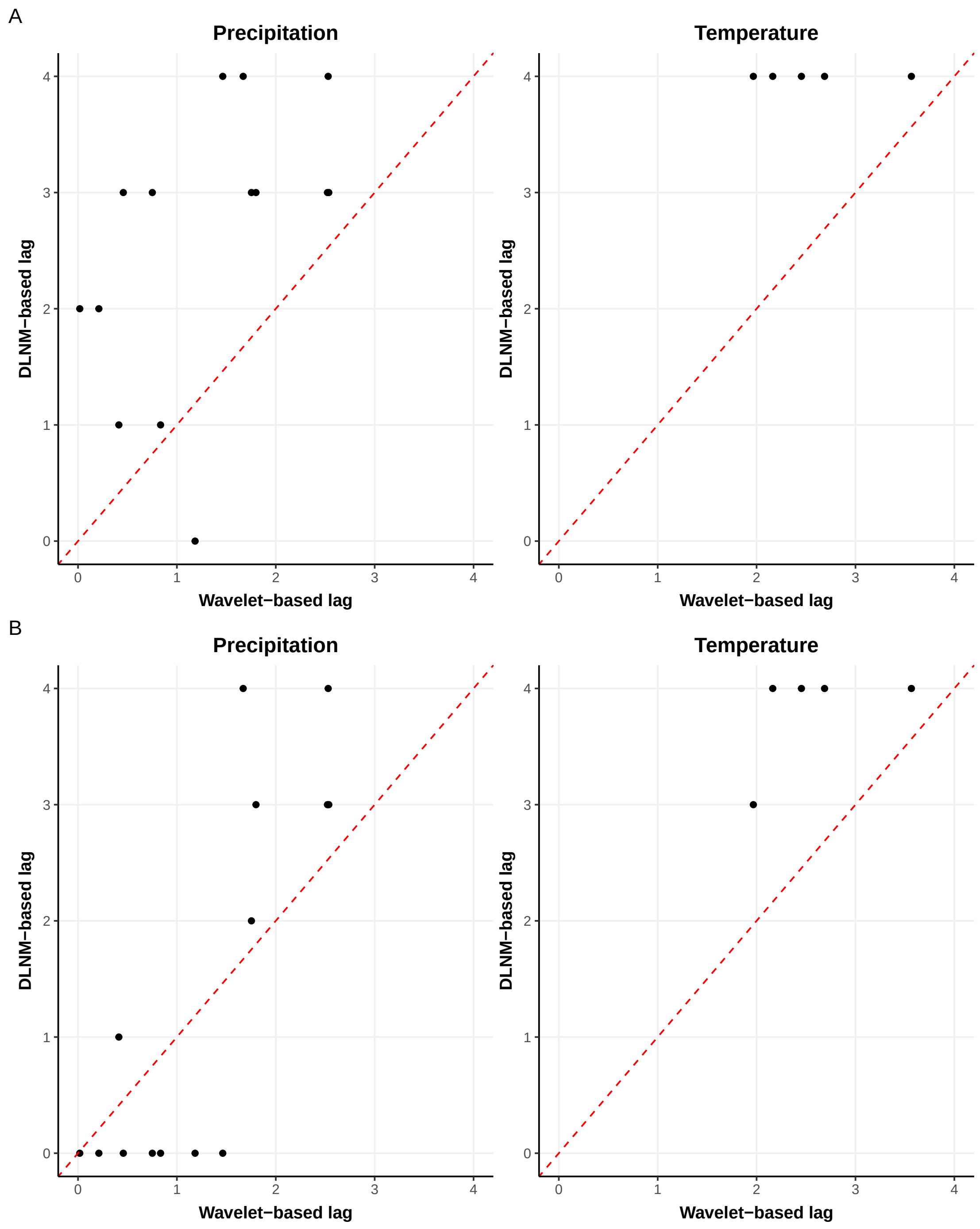

Figure 3: Comparison between wavelet-derived phase lags and DLNM-based optimal lags for precipitation and temperature. Panel A shows DLNM lags selected using the 'highest-risk' approach (lag with maximum cumulative RR); Panel B shows DLNM lags selected using the 'first-significant' approach (earliest lag with significant elevated risk). The red dashed line indicates perfect agreement. Divergence between methods is expected because wavelet phase assumes sinusoidal cycles while DLNM allows non-linear dose-response relationships.

#### 4.4 Figure S4: Year-by-Year Peak Month Variability (All Provinces)

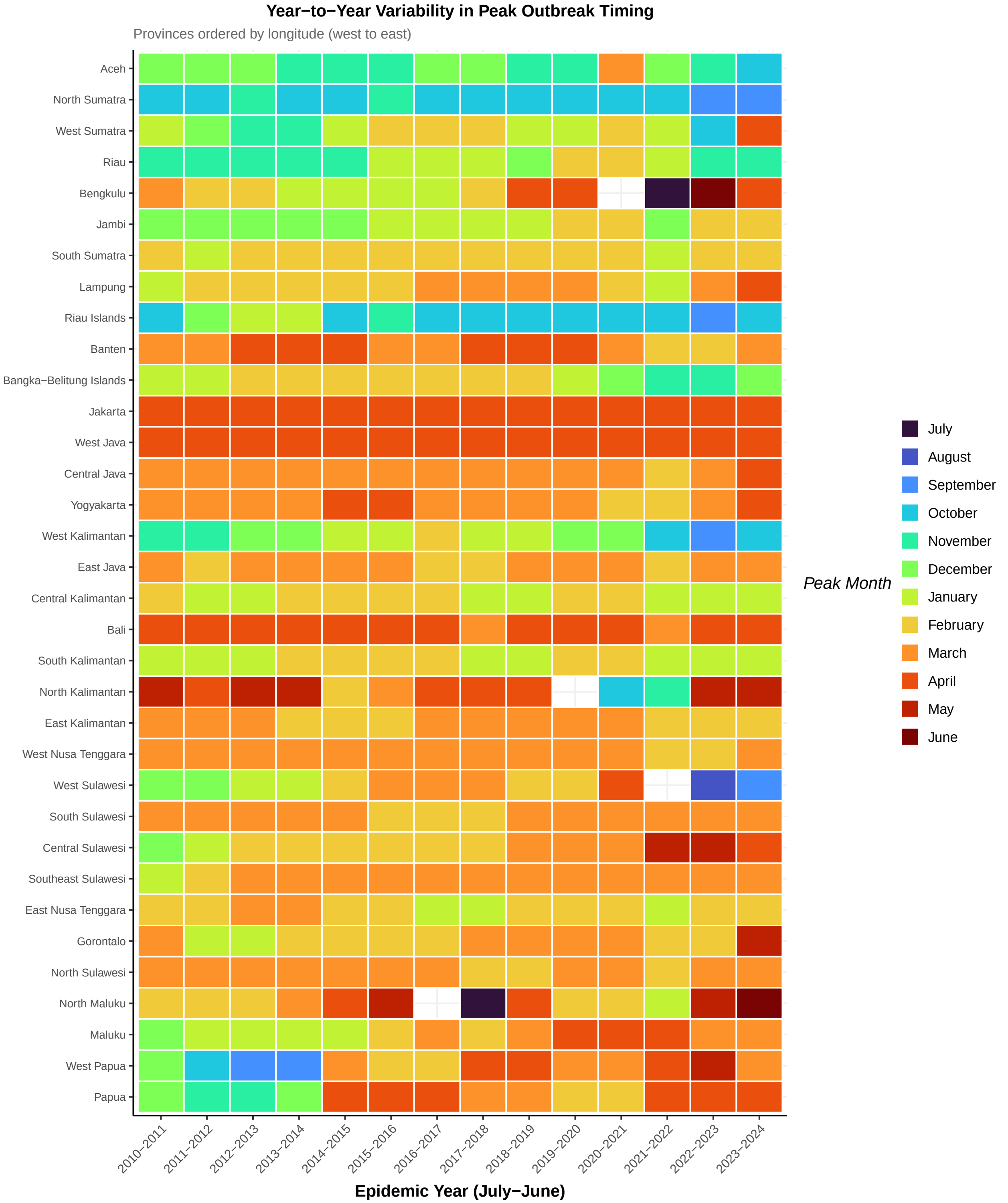

Figure 4: Year-to-year variability in peak outbreak timing across all 34 provinces. Provinces are ordered by longitude (west to east, bottom to top). Each cell shows the peak month for that province in that epidemic year (July-June). Grey cells indicate anomalous periods excluded due to weak annual periodicity. The heatmap reveals substantial inter-annual variability in peak timing, even within the same province, highlighting the complexity of dengue seasonality across Indonesia's archipelagic geography.

#### 4.5 Figure S5: Year-by-Year Peak Month Variability (Western Provinces)

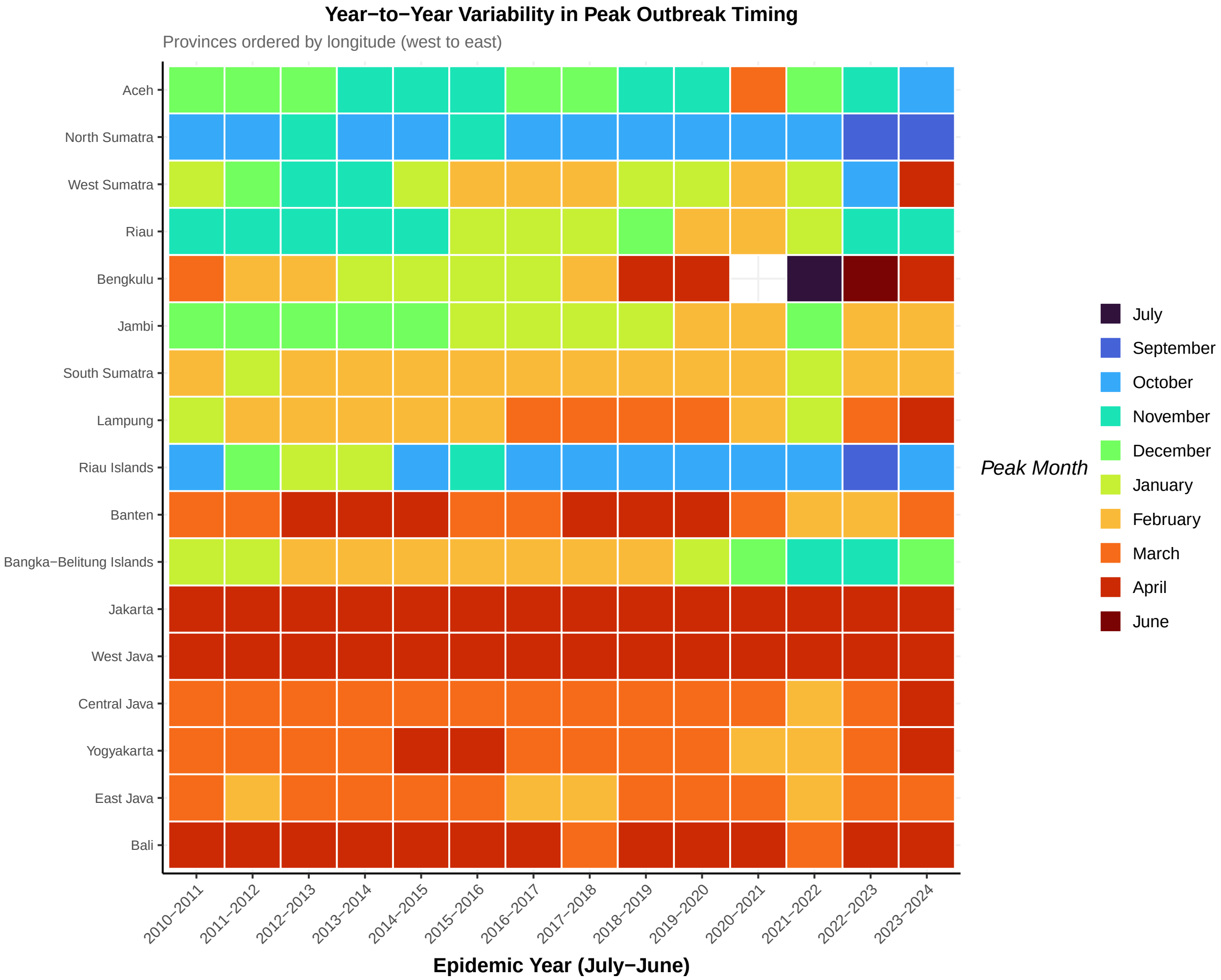

Figure 5: Year-to-year variability in peak outbreak timing for western provinces only (Sumatra and Java-Bali). Provinces are ordered by longitude (west to east, bottom to top). This subset shows the regions with clearest cluster structure (Figure 3C in main text). Despite stronger regional coherence than eastern provinces, substantial year-to-year variation in peak timing persists.
